# WHY A “NORMAL BLOOD VOLUME” IS NOT ALWAYS NORMAL – AN OVERLOOKED ISSUE IN HEART FAILURE MANAGEMENT

**DOI:** 10.64898/2026.08.18.26360762

**Authors:** Wayne L. Miller

**Affiliations:** Department of Cardiovascular Medicine, Mayo Clinic, Rochester, MN

**Author notes:** Corresponding Author: Wayne L. Miller, MD, PhD, FACC, FAHA, FHFSA, Mayo Clinic, Department of Cardiovascular Medicine 200 First Street, SW, Rochester, MN 55905.

**Keywords:** “Normal”Blood Volume, Chronic Heart Failure, RBC Mass, Plasma Volume, Heterogeneity in Blood Volume Profiles, Patient Management

## Abstract

**Background:** Blood volume (BV) in patients with chronic heart failure (HF) is characterized by heterogeneity in volume profiles; one profile being “normal BV”. While overall intravascular volume may be considered normal clinically, the relative contributions of red blood cell (RBC) mass and plasma volume (PV) may not be.

**Objective:** Assess how normal is a “normal BV” based on quantitative measures of RBC mass and PV.

**Methods:** Retrospective analysis was undertaken in 395 patients with Class II-III HF. BV was quantitated using indicator-dilution methodology. Cohort was stratified by normal and hypervolemic BV.

**Results:** Of the cohort, 31% (123/395) demonstrated normal total BV and 62% (244/395) hypervolemic BV. Of patients with “normal BV”, 36% (44/123) demonstrated normal RBC mass and 60% normal PV (74/123). Importantly, 60% (74/123) demonstrated a deficit in RBC mass (true anemia), while a low hemoglobin (<12 g/dL) was present in just 29% (36/123). An excess in RBC mass (erythrocytosis) in 4% (5/123). Notably, true normal BV (i.e., normal RBC mass and normal PV) was observed in only 30% (37/123) of patients with an overall “normal” intravascular volume.

**Conclusions:** Findings reveal that “normal BV” can be misleading by concealing substantial variability in RBC mass (including unrecognized anemia and erythrocytosis) as well as different degrees of PV expansion and contraction. An actual normal BV was identified in a minority of “normal BV” patients. This underscores the importance of looking beyond overall “normal BV” to the contributing elements of RBC mass and PV with significant implications for patient management and outcomes.

## Introduction

Clinically relevant heterogeneity in blood volume (BV) profiles has been identified in patients with stable chronic heart failure (HF) even when clinical features and patient presentations are similar (1-5). “Normal BV” is one profile contributing to this heterogeneity and is often equated to “euvolemia” as the desired guideline-recommended goal in the volume management of chronic HF (6). Rarerly, however, are differences in BV profiles taken into account, including normal BV, to guide fluid management. These profiles are subject to large variability in constituent elements of red blood cell (RBC) mass and plasma volume (PV) and which may themselves not always be normal. Further, in clinical practice a venous hemoglobin (Hb) concentration in the normal range (typically 12-17 g/dL) is considered to also reflect “normal BV” status. As a result, concluding that a total intravascular volume reported as normal or a hemoglobin concentration in the normal range accurately reflects a comprehensively normal BV can be misleading and conceal derrangements in RBC and PV and as a result missed opportunities to optimize treatment and improve outcomes.

Accordingly, the objectives of this analysis in a cohort of stable chronic HF patients were to determine 1) how normal is a “normal BV” profile when assessed by quantitative measures of RBC mass and PV; 2) how reliably is peripheral venous Hb concentrations concordant with “normal BV”; and 3) how do “normal BV” profiles compare in distribution of RBC mass and PV in patients with expanded BV profiles (hypervolemic) as contributors to the heterogeneity in intravascular volume and as factors influencing patient management.

## METHODS

### Study Group

Total BV and relevant clinical data were retrospecitvely analyzed from hemodynamically stable HF patients predominantly NYHA functional class II-III. BV was quantified using a standardized albumin-radiolabeled indicator-dilution methodology. Electronic medical records were reviewed and clinical laboratory test results were collected for all patients. Study exclusion criteria were 1) age <18 years; chronic kidney disease requiring hemodialysis or ultrafiltration; 3) symptomatic coronary artery disease; 4) patients requiring advanced pharmacologic or device therapy for hemodynamic support; 5) females who were pregnant or of child-bearing potential; 6) diagnosed congenital heart disease; 7) treatment resistant allergy to iodine which precluded BVA testing. Renal function was expressed as the estimated glomerular filtration rate (eGFR), mL/min/1.73m^2^, using the Modification of Diet in Renal Disease equation (7). Patients were receiving standard guide-line recommended HF medical therapy and were clinically stable during the period of evaluation and BV quantitative analysis. The Mayo Foundation Institutional Research Review Board and Ethics Committee approved the study protocol. All data are available in this report.

### Blood Volume Measurement

Total blood volume (BV) quantitation using the indicator-dilution method was undertaken in the Mayo Clinical Nuclear Medicine Laboratory by a standardized clinically available technique to administer low dose iodinated (I-131) labeled albumin intravenously (BVA-100 Blood Volume Analyzer/Volumex, Daxor Corp, Oak Ridge, TN). This is a computer-based technique for the direct measurement of plasma volume (PV). Reference normal volumes are defined by the deviation from ideal weight method (8-9). Normal volumes were determined from previously established sex-adjusted reference values adapted from life insurance tables as previously described (9-10). Each patient’s normal reference weight at his/her specific height was derived from the reference curves and percent deviation from normal expected weight was calculated as % expected weight = actual weight-expected normal weight/expected normal weight X 100. These data are electronically incorporated by algorithm into the BVA-100 analysis. The specifics of the technique and procedure have been previously reported (1,10-11)

Intravascular volumes are reported as absolute volumes (in liters) and as percent deviations from normal reference volume as deficit (-) or excess (+). Normal total BV, PV, and RBC mass were defined *a priori* as measured volume within ±8% (for total BV) and ±10% (for PV and RBC mass) of the individual’s expected normal volumes (9-11). As with previous reports definitions were the following: BV deficit (<-8% of normal BV), normal BV (≥-8% to ≤+8%), and BV expansion was defined as >+8% of normal volume. [Mild to moderate volume expansion (>+8% to ≤+25%) and large volume expansion (>+25% above normal)] (8,10). Based on World Health Organization (W.H.O.) criteria defining anemia (12), a low Hb (less than normal range) for the purposes of this analysis was established as a peripheral venous Hb of <12 g/dL.

### Statistical Analyses

Categorical variables are reported as number and percentage, while continuous variables are expressed as mean ± standard deviation (SD) if normally distributed or as median (25^th^-75^th^ interquartile range [IQR]) if non-normally distributed. To assess differences volume variables were compared using t-test for unpaired analyses, and percentage differences by Chi-squared analysis. Median values were compared using the Mann-Whitney test. Statistical analyses were performed using JMP and SAS version 9.3 with p-values <0.05 considered to be statistically significant.

## Results

Absolute BV for the cohort as a whole (N=395) was 6.3±1.7 (SD) liters (a 14% expansion above referenced normal BV). Of the cohort, 31% (123/395) demonstrated by quantitative analysis an overall “normal BV” and 62% (244/395) an expanded hypervolemic BV (defined as >8% above reference normal BV). Clinical characteristics, central hemodynamics and BV profiles are shown in Table 1. Of the patients with “normal BV”, 36% (44/123) demonstrated a normal RBC mass and 60% (74/123) a normal PV. The combination, however, of a normal RBC mass with a normal PV (true normal BV) was observed in just 30% (37/123) and for the cohort as a whole this was only 9.6% (38/395). Further, 60% (74/123) of the patients with “normal BV” demonstrated a deficit in RBC mass (true anemia defined as <-10% of referenced normal RBC mass volume). In contrast, a low Hb concentration of <12 g/dL, clinically consistent with W.H.O definition of anemia (12), was present in only 29% (36/123) providing a misleadingly low indicator of anemia compared with the actual RBC mass deficit in this cohort (a Hb-RBC mass mismatch,13-14). Dilutional pseudo-anemia [low Hb (<12 g/dL) but normal RBC mass with expanded PV (on average +10±4% above referenced normal PV)] was identified in 1.6% (2/123) of patiernts with “normal BV”. Additionallly, an excess in RBC mass (erythrocytosis, defined as >+10% above referenced normal RBC mass) was demonstrated in 4% (5/123) of patients with a “normal BV”. Figure 1 illustrates this heterogeneity of a “normal BV” profile.

**Table 1.** Clinical Characteristics, Hemodynamics and Volume Metrics in Patients with Chronic Heart Failure Stratified by Blood Volume Profiles.

| Variable | Normal BV<br>N=123 | Expanded BV<br>N=244 | P-Value |
| --- | --- | --- | --- |
| Age, years | 63±15 | 67±14 | 0.012 |
| Sex, M/F (% male) | 67/56 (54%) | 182/62 (75%) |  |
| BMI, kg/m<sup>2</sup><br>% ≥ 30 kg/m<sup>2</sup> | 32±8<br>54% | 33±7<br>55% | 0.259 |
| LVEF, %<br>% LVEF ≥50% | 45±17<br>46% | 37±17<br>29% | <0.001 |
| Blood Pressure, mm Hg,<br>systolic | 119±20 | 117±19 | 0.366 |
| Hemoglobin, g/dL<br>% < 12.0 g/dL | 12.9±2.0<br>29% | 12.4±2.2<br>40% | 0.035 |
| Hematocrit, % | 37±5 | 37±6 | 1.000 |
| GFR, mL/min/1.73m<sup>2</sup> | 57±22 | 52±21 | 0.036 |
| Plasma Albumin, g/dL | 4.1±0.5 | 3.9±0.5 | <0.001 |
| RAP, mmHg | 9±5 | 13±7 | <0.001 |
| PCWP, mmHg | 14±6 | 19±7 | <0.001 |
| mPAP, mmHg | 25±9 | 32±10 | <0.001 |
| TPG, mmHg | 5±4 | 7±6 | 0.042 |
| **Blood Volume, L** | 5.2±0.9 | 7.1±1.5 | <0.001 |
| % Excess (+)/Deficit (-)<br>from Normal Volume | +0.8±4%<br>+1 (-2, +5)% | +31±19%<br>+27 (+15, +43)% |  |
| % w/ Normal RBC Mass | 36% | 37% |  |
| % w/ RBC Mass Deficit | 60% | 17% |  |
| % w/ RBC Mass Excess | 4% | 46% |  |
| % w/ Normal PV | 60% | 4% |  |
| % w/ PV Expansion | 39% | 96% |  |
| **Plasma Volume, L** | 3.4±0.6 | 4.7±1.1 | <0.001 |
| % Excess (+)/Deficit (-)<br>from Normal Volume | +9±10%<br>+8 (+3, +16)% | +44±26%<br>+38 (+24, +60)% |  |
| **RBC Mass, L** | 1.8±0.4 | 2.4±0.7 | <0.001 |
| % Excess (+)/Deficit (-)<br>from Normal Volume | -12±13%<br>-13 (-21, -3)% | +12±24%<br>+8 (-5, +25)% |  |

Data expressed as Mean ± Standard Deviation, median (IQR 25%-75%); BV – blood volume; PV – plasma volume; RBC – red blood cell; GFR – glomerular filtration rate; BMI – body mass index; LVEF – left ventricular ejection fraction; Normal blood volume is defined as a range of -8% to +8% of expected normal reference volume; Expanded blood volume is defined as >+8% above expected normal reference volume.

**Figure 1.**
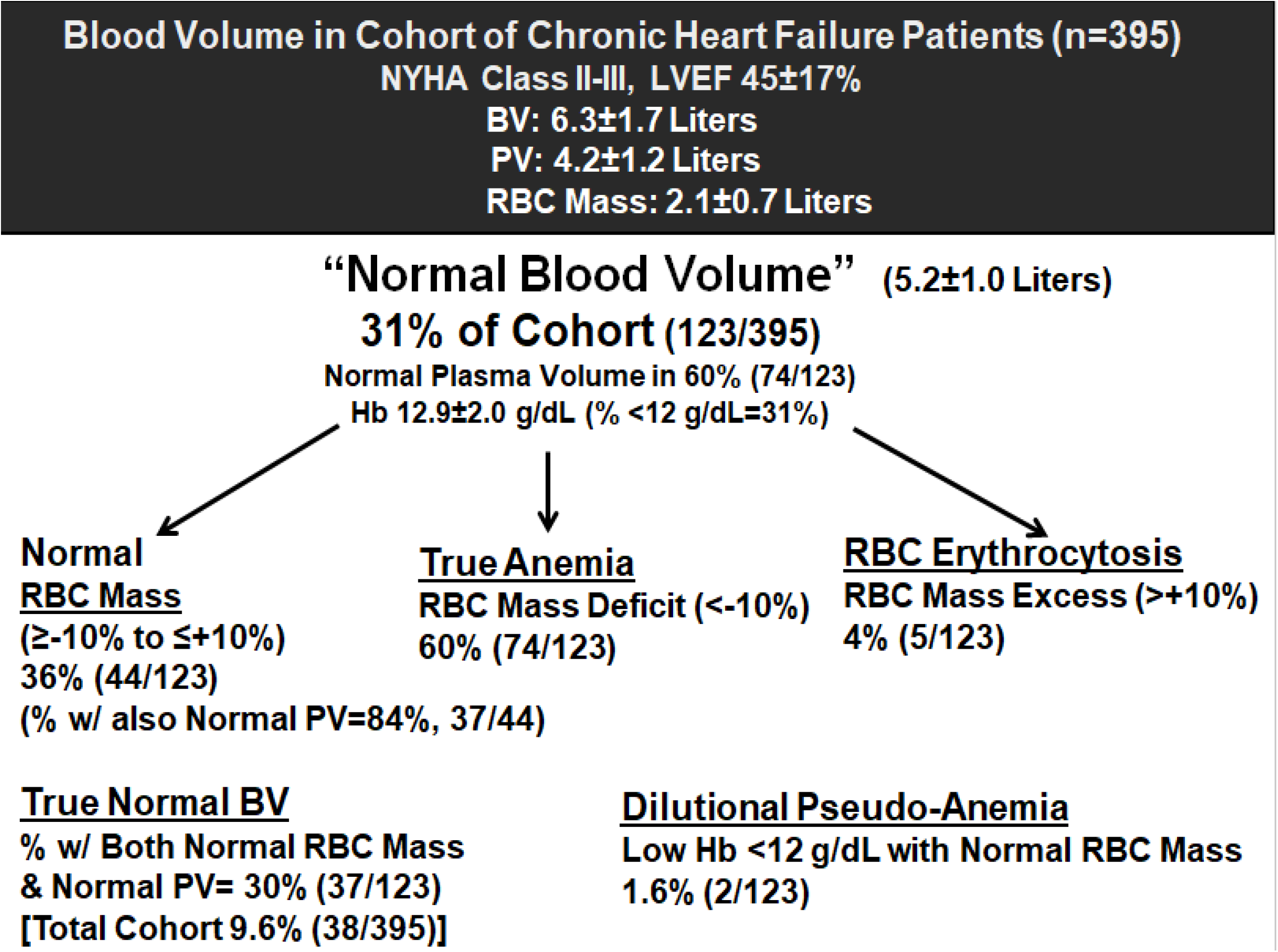
Composition of “Normal Blood Volume” Profile in a Cohort of Patients with Chronic Heart Failure.

Table 1 also provides data for the portion of this cohort with an expanded BV (244/395) which comprises the majority volume profile at 62%. RBC mass excess (erythrocytosis) was a large contributor to the BV expansion at 46% (112/244 vs. 4% in the “normal BV” profile) as was PV expansion (defined as PV >+10% of normal reference PV) at 96% (234/244 vs. 39% in the “normal BV” profile). RBC mass deficit (true anemia) was present in only 17% (41/244). Compared with “normal BV” patients, this subgroup was slightly older with a higher male prevalence, and lower LVEF. Hemoglobin, albumin and eGFR were also statistically different but only marginally lower from a clinical perspective. Figure 2 illustrates is contributing volume components comprising the expanded BV profile.

**Figure 2.**
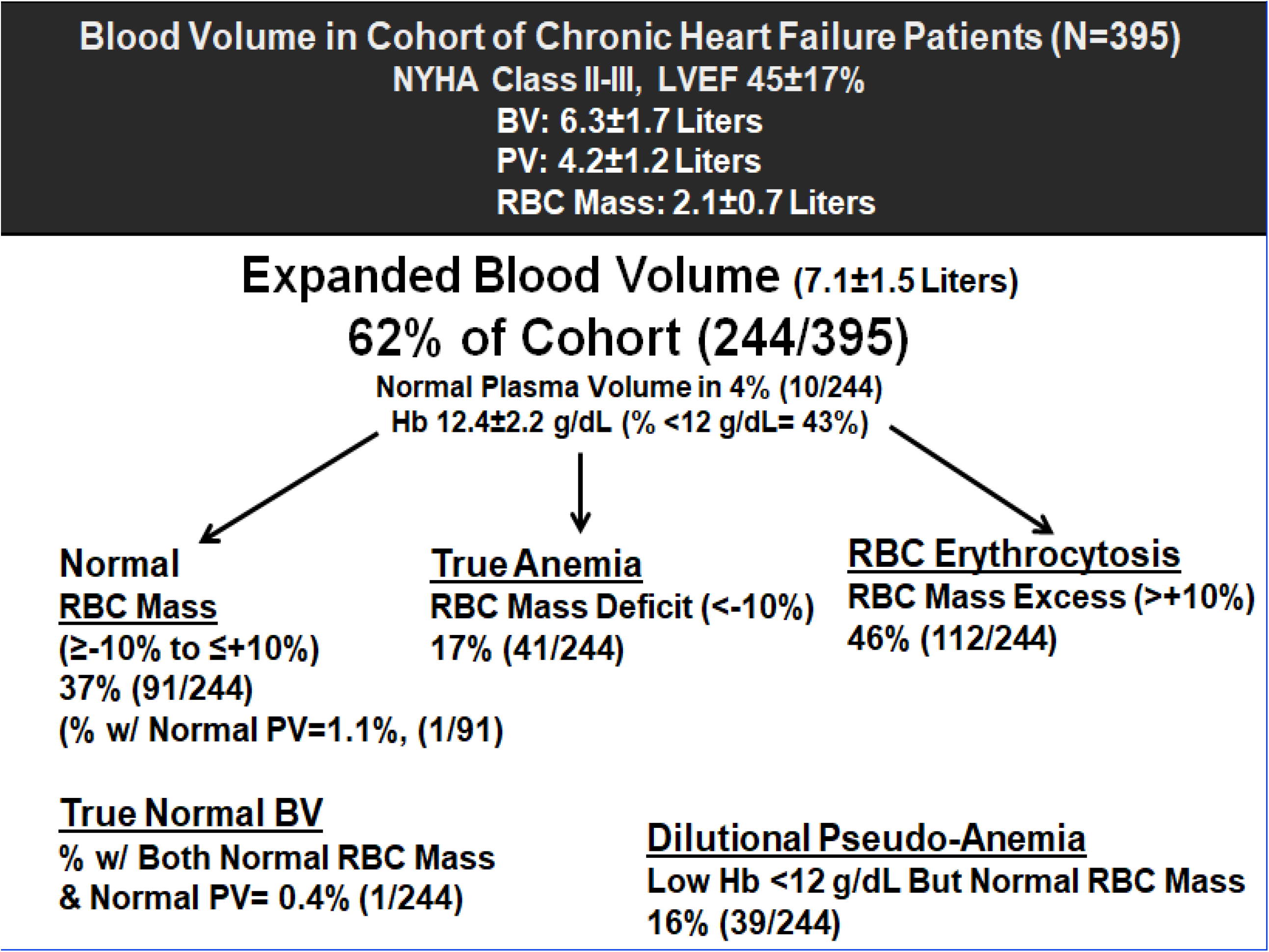
Composition of Expanded Blood Volume Profile in a Cohort of Patients with Chronic Heart Failure.

A subset of the patient cohort (normal BV, N=48; expanded BV, N=84) underwent clinically indicated right heart catheterization as part of their HF evaluation and provided hemodynamic data as also shown in Table 1. All hemodynamic metrics were higher and statistically different in the hypervolemic group relative to the normal BV group.

## Discussion

The results of this analysis in a large cohort of stable, chronic HF patients support the principle observation that a “normal BV” is actually normal in only a minority of patients. Four main findings are the following: 1) Significantly, a truly normal BV (i.e., normal RBC mass with normal PV) was observed in only a third of patients with a quantitatively total normal intravascular volume; 2) true anemia (deficit in RBC mass) was common in patients with an overall “normal BV” (60%) where PV was expanded to maintain an overall normal intravascular volume. Pseudo-anemia (dilutionally low hemoglobin concentration with normal or excess RBC mass) is also identifed in small percentage of patients with “normal BV” where PV has expanded to maintain an overall normal intravascular volume; 3) “Normal BV” profiles are also composed of contributions of RBC mass excess (absolute erythrocytosis) where PV is contracted to maintain an overall normal intravascular volume; and 4) an overall quantitatively normal BV was identifed in a minority of the patient cohort with chronic HF (approximately a third of the cohort) while the majority demonstrated an expansion in BV (hypervolemia) driven by RBC mass excess (erythrocytosis) and PV expansion.

These findings reflect that a “normal BV” is commonly not normal in terms of the individual contributions of RBC mass and PV which can vary significantly in their proportions, and frequently are not in normal range individually. This underscores the importance of understanding the marked variability in BV profiles (4,15-18) in patients with chronic HF in that while total intravascular volume may be normal overall, it can conceal true anemia, erythrocytosis, dilutional pseudo-anemia, as well as different degrees of PV expansion and contraction, all of which require different approaches to management in order to effectively guide appropriate treatment and also avoid unnecessary interventions.

One aspect of “normal BV” as identified in this analysis warrants particular attention and that is the high prevalence of true anemia (deficit in RBC mass) in HF patients with “normal BV”. Anemia is associated with poorer HF outcomes relative to normal RBC mass or even an excess in RBC mass (19-21). Also relevant is the detection of low venous Hb concentrations suggesting the presence of anemia, however, without also knowing RBC mass and PV, Hb values may provide misleading data by falsely suggesting anemia when PV has actually expanded and RBC mass, not a concentration metric, is actually normal. As a result dilutional pseudo-anemia (14, 22-23) may be misinterpreted and inappropriately intervened upon. Additionally, a normal range Hb may underestimate the degree of true anemia secondary to a contraction in PV. Therefore, understanding the variability in RBC mass and PV and their combinations even when overall BV is within a normal range can help guide the most appropriate management. This can be particulary relevant given the high prevalence of true anemia associated with “normal BV” and its implications for poor outcomes in patients with chronic HF. These factors all support the recommendation that “normal BV” be fully evaluated in patients with chronic HF where results carry significant implications for patient management and potentially clinical outcomes.

## Limitations

The retrospective design from a single center may not account for potential unidentified confounders contributing to differences in the distribution of volume profiles. The clinical significance of differences in BV profiles and their distributions, patient to patient, and across different HF cohorts requires further study.

## Conclusions

Findings reveal that the description of an overall “normal BV” can be misleading by concealing substantial variability in RBC mass and PV associated with the development of true anemia, absolute erythrocytosis as well as different degrees of PV expansion and contraction. An actual normal BV (normal RBC mass with normal PV) was identified in a minority (30%) of patients with a “normal BV”, while true anemia (absolute deficit in RBC mass) was present in 60% of “normal BV” patients. This underscores the importance of looking further into “normal total BV” to the composite elements of RBC mass and PV and the associated implications for more individualized patient management and impact on outcomes. Therefore, understanding that a reported “normal intravascular volume” may not be physiologically normal is of key importance in the management of patients with chronic HF.

## Data Availability

All data are included in the manuscript and tabled data.

## Clinical Perspectives

Findings of this analysis reveal that the clinical declaration of “euvolemia” or a “normal BV” can be misleading by concealing substantial variability in RBC mass and PV associated with true anemia, erythrocytosis as well as different degrees of PV expansion and contraction. An actual normal BV was identified in a minority (30%) of “normal BV” patients in the cohort studied. This underscores the importance of looking beyond “normal total BV” to the contributing elements of RBC mass and PV which carries significant implications for individualized patient management and outcomes.

## Notes

Disclosures: Unrestricted Research Grant Support by Daxor Corp., Oakridge, TN and the Feldschuh Foundation for Clinical Research, New Yotk, NY

No conflicts of interest to report

### Competing Interest Statement

The authors have declared no competing interest.

### Clinical Trial

Not a clinical trial

### Author Declarations

The Mayo Foundation Institutional Research Review Board and Ethics Committee approved the study protocol.

## References

1. Miller WL, Mullan BP. Understanding the heterogeneity in volume overload and fluid distribution in decompensated heart failure is key to optimal volume management: Role for blood volume quantitation. JACC Heart Fail 2014; 2: 298–305. doi: 10.1016/j.jchf.2014.02.007

2. Androne AS, Hryniewicz K, Hudaihed A, Mancini D, Lamanca J, Katz SD. Relation of unrecognized hypervolemia in chronic heart failure to clinical status, hemodynamics, and patient outcomes. Am J Cardiol 2004; 93:1254–1259.

3. Schuster C-J, Weil MH, Besso J, Carpio M, Henning RJ. Blood volume following diuresis induced by furosemide. Am J Med 1984; 75: 585–592.

4. Miller WL, Fudim M, Kittipibul V, Yaranov DM, Carry B, Silver MA. Understanding the Variability in Combinations of Red Cell and Plasma Volume Can Help Guide Management in Heart Failure. Eur Soc Cardiol Heart Fail 2025; 12: 142–149; epub 9-2024. doi: 10.1002/ehf2.15070.

5. Fudim M, Kittipibul V, Molinger J, Yaranov DM, Miller WL. Patient sex impacts volume phenotypes and hemodynamics in chronic heart failure –A multi-center analysis. J Cardiac Failure, 2025; 31: 379–287. 10.1016/j.cardfail.2024.05.013.

6. Maddox TM, Januzzi JL Jr, Allen LA, Breathett K, Brouse S, Butler J, Davis LL, Fonarow GC, Ibrahim NE, Lindenfeld J, Masoudi FA, Motiwala SR, Oliveros E, Walsh MN, Wasserman A, Yancy CW, Youmans QR. 2024 ACC Expert Consensus Decision Pathway for Treatment of Heart Failure With Reduced Ejection Fraction: A Report of the American College of Cardiology Solution Set Oversight Committee. J Am Coll Cardiol. 2024 Apr 16;83(15):1444–1488. doi: 10.1016/j.jacc.2023.12.024. Epub 2024 Mar 8. PMID: 38466244.

7. Levey AS, Coresh J, Balk E, Kausz AT, Levin A, Steffes MW, et al. National kidney foundation practice guidelines for chronic kidney disease: Evaluation, classification, and stratification. Ann Intern Med. 2003; 139:137–147. doi: 10.7326/0003-4819-139-2-200307150-00013.

8. Feldschuh J, Enson Y. Prediction of normal blood volume. Circulation. 1977; 56:605–615. 10.1161/01.CIRC.56.4.605

9. Feldschuh J, Katz S. The importance of correct norms in blood volume measurement. Am J Med Sci. 2007; 334:41–46. 10.1097/MAJ.0b013e318063c707.

10. Feldschuh J. Blood volume measurements in hypertensive disease. In: Larah JK, Brenner BM, editors. Hypertension: Pathology, Diagnosis, and Management. New York: Raven Press, 1990.

11. Miller WL. Measurement of blood volume in patients with heart failure: Clinical relevance, surrogates, historical background and contemporary methodology. Heart International Journal. 2023; 17: 36–44. doi.org/10.17925/HI.2023.17.1.36.

12. World Health Organization. Hemoglobin concentration for the diagnosis of anemia and assessment of severity 2011. Who.int/vmnis/indicators/haemoglobin/en/.

13. Miller WL, Mullan BP. Peripheral venous hemoglobin and red blood cell mass mismatch in volume overload systolic heart failure: Implications for patient management. J Cardiovasc Transl Res 2015; 8: 404–410. doi: 10.1007/s12265-015-9650-4.

14. Otto JM, Plumb JOM, Clissold E, Kumar SB, Wakeham DJ, Schmidt W, et al. Hemoglobin concentration, total hemoglobin mass and plasma volume in patients: Implications for anemia. Haematologica 2017; 102: 1477–1485. doi.10.3324/heamatol.2017.169680.

15. Miller WL, Grill DE, Mullan BP. Differences in red blood cell mass profiles impact intravascular volume and outcome risk in chronic heart failure. Eur Soc Cardiol - Heart Failure 2023; 10: 1270–1279; epub January 30, 2023. doi: 10.10002/ehf2.14309.

16. Nust P, Verbrugge FH, Bertrand PB, Martens P, Dupont M, Drieskens O, et al. Plasma volume is normal but heterogeneously distributed, and true anemia is highly prevalent in patients with stable heart failure. J Card Fail 2017; 23: 138–144. 10.1016/j.cardfail.2016.08.008.

17. Ahlgrim C, Birkner P, Seiler F, Wrobel N, Grundmann S, Bode C Pottgiesser. Increased red cell volume is a relevant contributing factor to an expanded blood volume in compensated systolic heart failure. J Card Fail 2020; 26: 420–428. doi: 10.106/j.cardfail.2019.11.025.

18. Miller WL, Grill DE, Mullan BP. Comparison of blood volume profiles in heart failure with preserved and reduced ejection fractions – Sex makes a difference. Circulation-Heart Failure 2024 17: e010906, doi: 10.1161/CIRCHEARETFAILURE.123.010609.

19. Ezekowitz JA, McAlister FA, Armstrong PW. Anemia is common in heart failure and is associated with poor outcomes. Insights from a cohort of 12065 patients with new onset heart failure. Circulation 2003; 107: 223–225. doi.org/10.1161/01.CIR.0000052622.FC

20. Miller WL, Grill DE, Mullan BP. Differences in red blood cell mass profiles impact intravascular volume and outcome risk in chronic heart failure. Eur Soc Cardiol - Heart Failure 2023; 10: 1270–1279; epub January 30, 2023. doi. 10.10002/ehf2.14309.

21. Miller WL, Strobeck JM, Grill DM, Mullan BP. Blood volume expansion, normovolemia and clinical outcomes in chronic human heart failure. Am J Physiol: Heart & Circ Physiol 2021; 32: H1074 H1082. doi: 10.1152/ajpheart.00336.2021.

22. Androne A-S, Katz SD, Lund L, LaManca J, Hudaihed A, Hryniewicz K, Mancini DM. Hemodilatuion is common in patients with advanced heart failure. Circulation 2003; 107: 226–229. doi: 19.1161/01.CIR.0000052623.16194.80.

23. Van PY, Riha GM, Cho D, Underwood SJ, Hamilton GJ, Anderson R, Ham B, Schreiber MA. Blood volume analysis can distinguish true anemia from hemodilution in critically ill patients. J Trauma 2011; 70: 646–651. doi:10.1097/TA.06013e1820d5f48.

